# Fetal and Neonatal Echocardiographic Analysis of Biomechanical Alterations for the Hypoplastic Left Heart

**DOI:** 10.1101/2020.10.21.20217265

**Authors:** Brett A. Meyers, Sayantan Bhattacharya, Melissa C. Brindise, Yue-Hin Loke, R. Mark Payne, Pavlos P. Vlachos

**Author notes:** Corresponding Author: Pavlos P. Vlachos, 585 Purdue Mall, West Lafayette, IN, 47907, Fax: *none. d. Tweet: NEW ARTICLE IN #JACCIMG. New echocardiography tools advance our understanding of congenital heart disease in the fetus. #cvImaging #PedsCards #CHD #CardioTwitter.

## Abstract

**Background:** Hypoplastic left heart syndrome (HLHS) presents diagnostic and prognostic challenges while progressing toward heart failure (HF). Understanding the fetal and neonatal HLHS biomechanics, including novel hydrodynamic parameters, could help better planning of the long-term management of HLHS patients.

**Objectives:** Compare fetal and neonatal HLHS cardiac biomechanics against normal subjects using echocardiography.

**Methods:** We performed a retrospective study of 10 HLHS patients with echocardiograms at 33-weeks gestation and at the first week post-birth and 12 age-matched controls. We used in-house developed analysis algorithms to quantify ventricular biomechanics from four-chamber B-mode and color Doppler scans. Cardiac morphology, hemodynamics, tissue motion, deformation, and flow parameters were measured.

**Results:** Tissue motion, deformation, and index measurements did not reliably capture biomechanical changes. Stroke volume and cardiac output were nearly twice as large for the HLHS right ventricle (RV) compared to the control RV and left ventricle (LV) due to RV enlargement. The enlarged RV exhibited disordered flow with higher energy loss (EL) compared to prenatal control LV and postnatal control RV and LV. Furthermore, the enlarged RV demonstrated elevated vortex strength (VS) compared to both the control RV and LV, prenatally and postnatally. The HLHS RV showed reduced relaxation with increased early filling velocity (*E*) compared prenatally to the LV and postnatally to the control RV and LV. Furthermore, increased recovery pressure (Δ*P*) was observed between the HLHS RV and control RV and LV, prenatally and postnatally.

**Conclusions:** The novel hydrodynamic parameters more reliably capture the HLHS alterations in contrast to traditional parameters.

## Introduction

Hypoplastic left heart syndrome (HLHS) is a congenital heart defect that affects 1 in ever 4,000 births (1,2), with poor survival (3) and high annual treatment costs (4). Detecting HLHS via fetal ultrasound enables earlier care planning which improves outcomes and costs (5), though accurate diagnosis *in utero* does present challenges (6). Fetal echocardiography is recommended to guide these high-risk pregnancies (7), and pediatric echocardiography helps guide treatment from birth through adolescence (8). These exams routinely collect systolic function measurements [i.e., stroke volume (SV), cardiac output (CO)] which are often inaccurate, highly variable, and affected by image quality, probe placement, heart size, and tissue motion (7,9). Furthermore, there is a lack of clinical understanding on assessment of diastolic function in the right ventricle (RV), which is significantly altered in the perinatal period but important for HLHS physiology (10,11).

Fetal echocardiography can potentially provide relevant diastolic function parameters which includes speckle tracking strain (7,12) and hemodynamics (9). However, clinical acceptance of fetal strain measurement lags as commercial software are vendor-specific (13), have strict application requirements (14), and requires expert training (15). Moreover, these software rely on chamber segmentation, which is uncommon in fetal echocardiography due to poor image quality and model assumptions (7). When performed, segmentations are hand-drawn and undergo correction, which increases observer variability and user time. Color Flow Imaging (CFI) is used clinically to detect septal and valve defects; however, flow patterns can also be observed. With the help of quantitative tools, flow-induced vortices, energy losses, and pressure distributions can be resolved which can help characterize the abnormal flow patterns present in HLHS hearts (8).

This study applies an integrated and automated echocardiography analysis method for measuring cardiac biomechanics from fetal and neonatal echocardiograms. The analysis collects chamber, annular motion, strain, and hydrodynamics parameters. The employed tools are not based on machine learning or shape models, are vendor-agnostic, and do not rely on heuristics adopted from adult echocardiography. These advancements enable conventional and novel biomechanics measurements to be robustly collected from fetal and neonatal echocardiograms.

Our study explores how biomechanics parameters quantified from echocardiography differ between the healthy left (LV) and right ventricle (RV), and HLHS RVs for two-time points, during gestation and day of birth. We hypothesize that HLHS RV has altered diastolic flow compared to both sides of the healthy heart, captured by the biomechanics parameters. The HLHS presents a demonstratively challenging clinical scenario that punctuates the need for robust parameters to comprehensively understand patient cardiac health.

## Methods

### Study population

Patient examinations were retrospectively selected from within the Indiana University Health and Children’s National Hospital networks. The cohort comprised ten HLHS subjects with fetal echocardiography performed at 33 weeks average gestational age and with pediatric transthoracic echocardiography at day of birth. Twelve age-matched healthy controls were included. Datasets without B-mode and CFI recordings in the apical long-axis (ALAX) view were excluded. All exams were deidentified prior to the data sharing between institutions for analysis. The Institutional Review Board for Human Studies for all institutions approved the study.

### Echocardiography

Sonographers performed fetal and pediatric echocardiograms on one of either Acuson SC200 ultrasound systems (Siemens Medical Solutions USA, Inc., Malvern, Pennsylvania), iE33/Epic 7 (Philips, Andover, Massachusetts), or Vivid E-95 (General Electric, Boston, MA, USA) ultrasounds. American Society of Echocardiography guidelines were followed (8,9). ALAX B-mode and CFI acquisitions were obtained with appropriate Nyquist limit and color box covering the entire ventricular cavity.

### Image analysis workflow

The analysis workflow, summarized in Figure 1, outputs ventricular cardiac biomechanics measurements from B-mode and CFI ALAX recordings. These modalities are utilized because sonographers are well-trained in their recording, which enhances analysis consistency. The workflow automates measurements, enabling once challenging and highly user-variable analysis to become routine. All algorithms were run in MATLAB (The MathWorks, Natick, Massachusetts).

**Figure 1:**
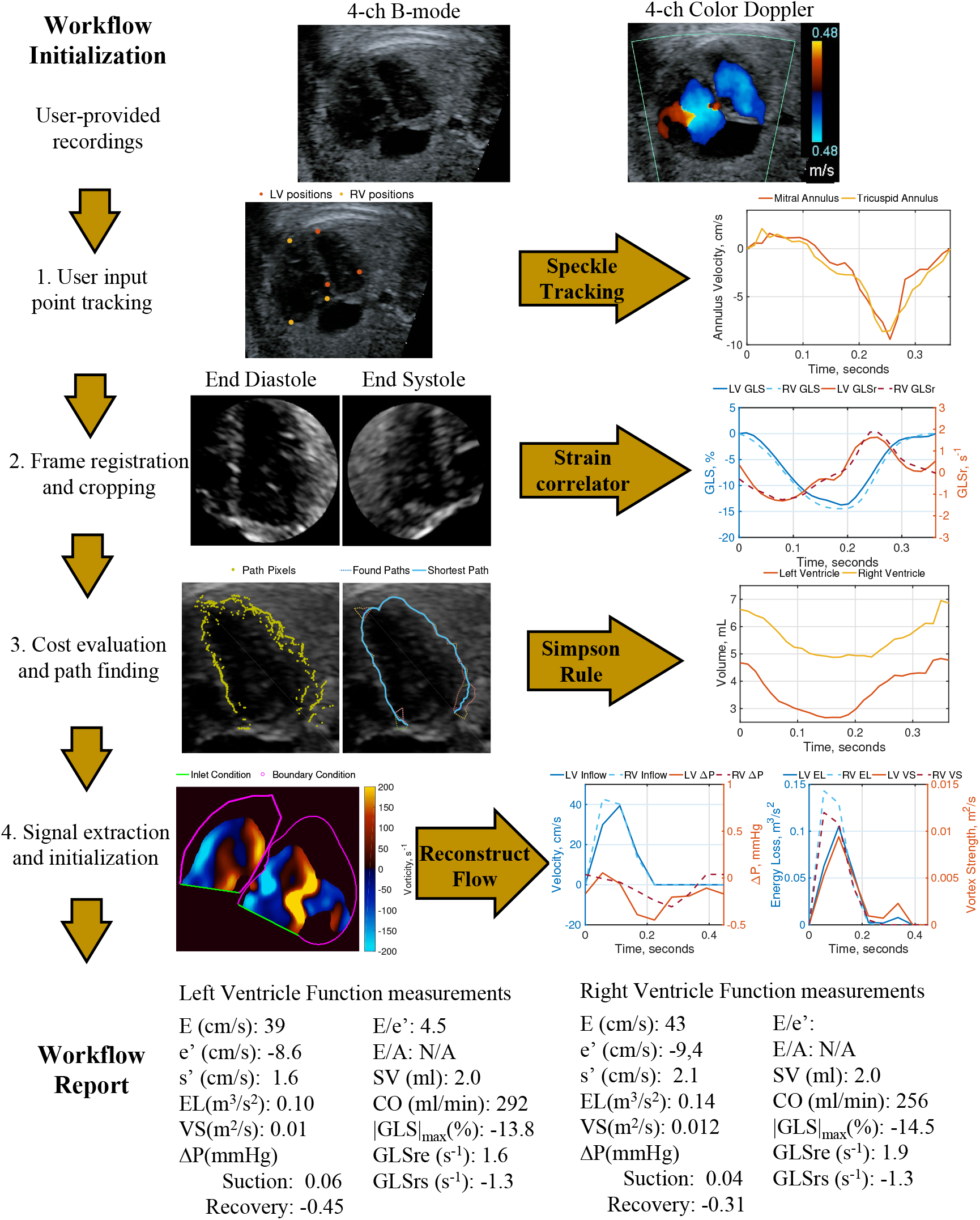
Echocardiogram analysis workflow. Analysis begins with the user providing 4C views. (1) AV annulus and apex feature points are provided to initialize automated analysis. (2) B-mode frames are co-registered, cropped, and processed to quantify GLS. (3) B-mode frames are evaluated to find pixel costs and paths for ventricle segmentation and volume quantification. (4) Color Doppler frames are processed to extract the signal, segment the ventricle, set initial conditions, and reconstruct velocity fields. Cardiac function measurements are compiled into a workflow report.

#### Step 1: Tracking user input annulus and apex positions

One set of user inputs marking the ventricle apex and atrioventricular (AV) annulus positions on the first recorded frame, depicted in Figure 1-**1**, are required for each scan. These inputs are tracked temporally using a speckle tracking algorithm, described in Appendix A1. The tracked positions provide measurements of ventricle relaxation that occurs during diastole and contraction that occurs during systole (16), driven by the AV annulus.

AV annulus positions are differentiated temporally to obtain velocities and adjusted relative to the apex, which is assumed stationary during the heartbeat. Peak annulus velocities for systolic ejection (s’) and early diastolic filling (e’) are automatically measured. Automated speckle tracking mitral annulus position and velocity measurements has been previously validated (17).

#### Step 2: Global longitudinal strain

A novel algorithm is used to measure GLS from the whole ventricle image (18), bypassing the above limitations. Briefly, the B-mode recording frames are co-registered using the tracked positions and cropped to keep the ventricle image, shown in Figure 1-**2**. A specialized correlation kernel estimates GLS rate between frames, which are then integrated to resolve GLS. The kernel is described in Appendix A2. Peak GLS (|GLS|, max) is output to quantify deformation.

#### Step 3: Unsupervised chamber segmentation

The unsupervised segmentation tool (ProID) automates ventricle detection and volume estimation (19). The tool identifies ventricle boundaries using a machine vision algorithm (20) that finds the shortest path of pixels around the ventricle image, shown in Figure 1-**2**. ProID overcomes contrast-to-noise and resolution limitations common to the natal imaging (7) by employing an echocardiogram-specific cost-matrix. The tracked positions are used to initialize ProID for each frame. Further description is provided in Appendix A3. Segmentation-derived volumes are computed using Simpson rule. SV and CO are output which quantify systolic function.

#### Step 4: Color Flow Imaging hemodynamics analysis of diastolic flow

Doppler vector reconstruction (DoVeR) resolves the underlying 2D velocity vector field of blood flow within the ventricle from CFI using the relationship between flow rate and fluid rotation (21). DoVeR uses the tracked positions and ProID to segment the ventricle in each frame. These segmentations are used to set boundary conditions for the DoVeR algorithm, shown in Figure 1-**3**. The vector fields are evaluated for peak early filling velocities (E), energy loss (EL), and vortex strength (VS) as well as the annulus-to-apex recovery pressure difference (recovery Δ*P*; IVPD) and AV valve center to minimum pressure distance (AV-to-P_min_) from computed pressure fields. Dimensionless quantities for E/e’ and E/A were also computed. Only diastolic flow assessment was emphasized, as the traditional apical view does not adequately profile the outflow in the imaging plane of the transducer. Further description of DoVeR and pressure field reconstruction are provided in Appendix A4.

### Statistical Analysis

Data are reported as the mean with 95% confidence intervals. We compared each parameter across conditions using the paired Student’s T-test. A two-tailed p-value < 0.05 was considered statistically significant. We performed statistical analysis using the MATLAB Statistics toolbox. Additional metrics computed but not presented are provided in Appendix C.

## Results

### Subject demographics

Relevant clinical information for each of the 10 HLHS subjects are provided in Table 1. The cohort composed of 6 males and 4 females. Mitral atresia was the most common subtype, affecting 5 patients, followed by mitral stenosis (3) and restrictive patent foramen ovale (PFO; 1). One subject did not have a reported subtype.

**Table 1:**
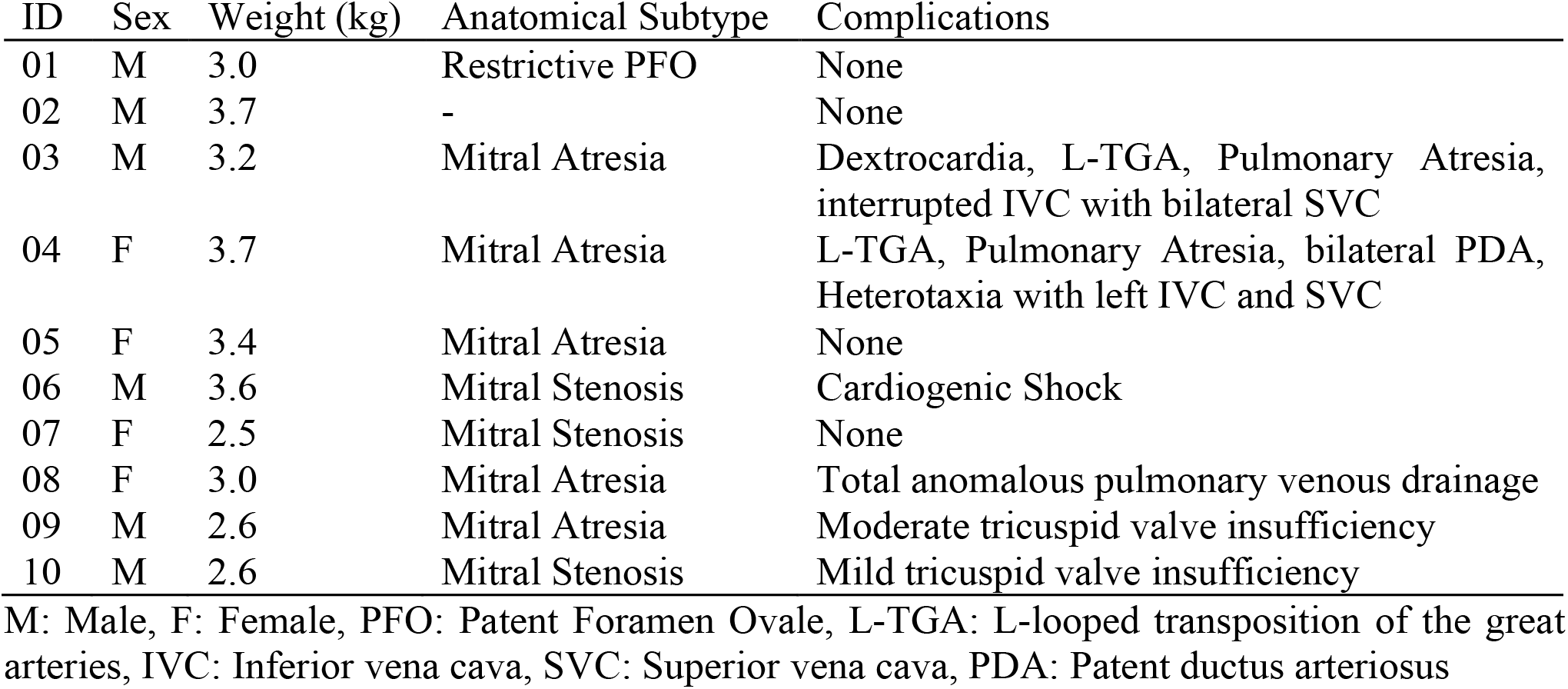
Demographics of hypoplastic left heart subjects used in this study

| ID | Sex | Weight (kg) | Anatomical Subtype | Complications |
| --- | --- | --- | --- | --- |
| 01 | M | 3.0 | Restrictive PFO | None |
| 02 | M | 3.7 | - | None |
| 03 | M | 3.2 | Mitral Atresia | Dextrocardia, L-TGA, Pulmonary Atresia, interrupted IVC with bilateral SVC |
| 04 | F | 3.7 | Mitral Atresia | L-TGA, Pulmonary Atresia, bilateral PDA, Heterotaxia with left IVC and SVC |
| 05 | F | 3.4 | Mitral Atresia | None |
| 06 | M | 3.6 | Mitral Stenosis | Cardiogenic Shock |
| 07 | F | 2.5 | Mitral Stenosis | None |
| 08 | F | 3.0 | Mitral Atresia | Total anomalous pulmonary venous drainage |
| 09 | M | 2.6 | Mitral Atresia | Moderate tricuspid valve insufficiency |
| 10 | M | 2.6 | Mitral Stenosis | Mild tricuspid valve insufficiency |

M: Male, F: Female, PFO: Patent Foramen Ovale, L-TGA: L-looped transposition of the great arteries, IVC: Inferior vena cava, SVC: Superior vena cava, PDA: Patent ductus arteriosus

### Systolic parameters in fetal HLHS RV vs. normal RV/LV

Measured parameters for fetal control and HLHS hearts are provided in Table 2. Major differences were observed for morphology but also for systolic parameters. SV (ml) and CO (ml/min) were elevated for the HLHS RV compared to the control RV and LV by a nearly two-fold statistically significant difference. Peak s’ (cm/s) was comparable for the HLHS RV and both control ventricles. Peak GLS for the HLHS RV was elevated compared to the control LV but not the RV.

**Table 2:** Echocardiographic measurements obtained from automated analysis platform

|  |  | HLHS RV (n = 10) | CTRL LV (n = 12) |  | CTRL RV (n = 12) |  |
| --- | --- | --- | --- | --- | --- | --- |
| | | $\mu \pm 1\sigma$ | $\mu \pm 1\sigma$ | p-value | $\mu \pm 1\sigma$ | p-value |
| **Systolic Parameters** |  |  |  |  |  |  |
| Stroke Volume<br>(ml) | Prenatal | 4.20 ± 2.08 | 1.85 ± 0.79 | **0.002** | 2.52 ± 1.18 | **0.030** |
| | Postnatal | 4.38 ± 2.54 | 3.23 ± 1.22 | 0.187 | 3.42 ± 3.05 | 0.455 |
|  | p-value | 0.868 | **0.004** |  | 0.366 |  |
| Cardiac Output<br>(ml/min) | Prenatal | 618 ± 324 | 282 ± 111 | **0.004** | 386 ± 183 | 0.052 |
| | Postnatal | 675 ± 530 | 426 ± 181 | 0.144 | 444 ± 429 | 0.283 |
|  | p-value | 0.771 | **0.034** |  | 0.683 |  |
| s',<br>(cm/s) | Prenatal | 2.24 ± 1.57 | 2.77 ± 1.02 | 0.365 | 3.37 ± 1.65 | 0.117 |
| | Postnatal | 3.17 ± 1.01 | 2.20 ± 0.81 | **0.021** | 3.00 ± 0.87 | 0.677 |
|  | p-value | 0.131 | 0.149 |  | 0.498 |  |
| \|GLS\|<sub>max</sub><br>(%) | Prenatal | 19.4 ± 6.3 | 15.5 ± 3.0 | 0.083 | 19.2 ± 10.0 | 0.966 |
| | Postnatal | 16.6 ± 3.6 | 19.3 ± 5.2 | 0.183 | 23.6 ± 8.2 | **0.023** |
|  | p-value | 0.249 | **0.045** |  | 0.265 |  |
| **Diastolic Parameters** |  |  |  |  |  |  |
| e'<br>(cm/s) | Prenatal | 3.63 ± 1.96 | 3.45 ± 1.07 | 0.798 | 4.68 ± 2.16 | 0.246 |
| | Postnatal | 4.22 ± 1.32 | 3.59 ± 1.85 | 0.374 | 3.93 ± 1.53 | 0.639 |
|  | p-value | 0.429 | 0.836 |  | 0.345 |  |
| E<br>(cm/s) | Prenatal | 42.1 ± 10.0 | 30.4 ± 6.5 | **0.004** | 35.0 ± 9.6 | 0.105 |
| | Postnatal | 62.0 ± 9.4 | 42.5 ± 17.5 | **0.006** | 34.6 ± 16.8 | **< 0.001** |
|  | p-value | **< 0.001** | **0.045** |  | 0.950 |  |
| E/e' | Prenatal | 16.2 ± 11.3 | 10.0 ± 4.6 | 0.108 | 9.5 ± 6.2 | 0.097 |
| | Postnatal | 16.2 ± 5.8 | 12.2 ± 5.4 | 0.122 | 10.1 ± 3.4 | **0.011** |
|  | p-value | 0.998 | 0.338 |  | 0.765 |  |
| E/A | Prenatal | 1.27 ± 0.39 | 1.36 ± 0.44 | 0.657 | 1.27 ± 0.55 | 0.962 |
| | Postnatal | 1.61 ± 0.44 | 1.40 ± 0.65 | 0.397 | 0.84 ± 0.27 | **< 0.001** |
|  | p-value | 0.082 | 0.859 |  | 0.066 |  |
| Flow energy loss<br>(mW) | Prenatal | 0.25 ± 0.22 | 0.09 ± 0.07 | **0.026** | 0.14 ± 0.11 | 0.149 |
| | Postnatal | 0.56 ± 0.49 | 0.19 ± 0.20 | **0.044** | 0.16 ± 0.19 | **0.030** |
|  | p-value | 0.077 | 0.103 |  | 0.756 |  |
| Vortex Strength<br>(cm<sup>2</sup>/s) | Prenatal | 241 ± 108 | 104 ± 42 | **< 0.001** | 146 ± 55 | **0.017** |
| | Postnatal | 322 ± 135 | 197 ± 110 | **0.035** | 155 ± 110 | **0.007** |
|  | p-value | 0.143 | **0.016** |  | 0.826 |  |
| Recovery ΔP<br>(mmHg) | Prenatal | -1.17 ± 0.81 | -0.50 ± 0.27 | **0.018** | -0.61 ± 0.22 | **0.039** |
| | Postnatal | -2.34 ± 1.13 | -1.10 ± 1.02 | **0.019** | -0.68 ± 0.63 | **< 0.001** |
|  | p-value | **0.013** | 0.075 |  | 0.707 |  |
| Min. ΔP Location<br>(mm) | Prenatal | 9.1 ± 3.6 | 4.6 ± 1.9 | **0.002** | 4.6 ± 1.7 | **0.001** |
| | Postnatal | 7.9 ± 2.8 | 3.6 ± 2.0 | **< 0.001** | 4.2 ± 2.1 | **0.003** |
|  | p-value | 0.419 | 0.254 |  | 0.599 |  |

s': Systolic annulus velocity, |GLS|<sub>max</sub>: Peak absolute global longitudinal strain, e': Early diastolic annulus velocity, E: Early diastolic filling velocity, E/e': Early diastolic annulus to filling velocity ratio, E/A: Early to late diastolic filling velocity ratio, ΔP: Pressure difference

### Diastolic parameters in fetal HLHS RV vs. normal RV/LV

Major differences were observed for several of the diastolic parameters. HLHS RV E (cm/s) was significantly elevated compared to the control RV and LV. The E/e’ quantity, an estimate of filling pressure (16), was elevated in the HLHS RV compared to the control RV and LV. Conversely, the E/A quantity, which helps identify diastolic dysfunction (16), was comparable for the HLHS RV against both the control ventricles. Peak flow EL (FEL), a measurement of total EL over the ventricle volume in the HLHS RV (mW) was elevated compared to the control RV and significantly different to the LV. Peak VS in the HLHS RV (cm^2^/s) was significantly elevated compared to both the control RV and LV. Recovery Δ*P* was significantly elevated in the HLHS RV compared to the control RV and LV. AV-to-P_min_ occurred significantly further from the annular plane for the HLHS RV compared to the control RV and LV.

### Qualitative assessment between fetal HLHS RV and normal controls

Representative changes in ventricular volumes, strains, and intracardiac flows are illustrated in Figure 2 for a fetal HLHS RV vs. fetal control. The HLHS RV exhibits several major differences compared to the control RV. First, the HLHS RV volume is larger (Figure 2a) and has an altered AV valve position, producing an asymmetric vortex pair during diastole (Figure 2c, d), with the free wall vortex occupying a larger area than the septal wall vortex. Second, an augmented pressure field was observed during both early diastole (Figure 2c-1), and late diastole (Figure 2c-2), where the free wall vortex had stronger low pressure and the apex had stronger high pressure compared to the healthy heart. Third, stronger EL was observed during both early diastole (Figure 2d-1) and late diastole (Figure 2d-2) compared to the healthy heart due to more disordered flow which produces greater shear. Fourth, the HLHS RV time-series (Figure 2c, d) did not show distinctly separate early and late diastolic filling; instead, these phases were fused. Thus, the HLHS fetal RV experienced stronger reversal IVPD and peak FEL compared to the healthy heart due to altered filling patterns.

**Figure 2:**
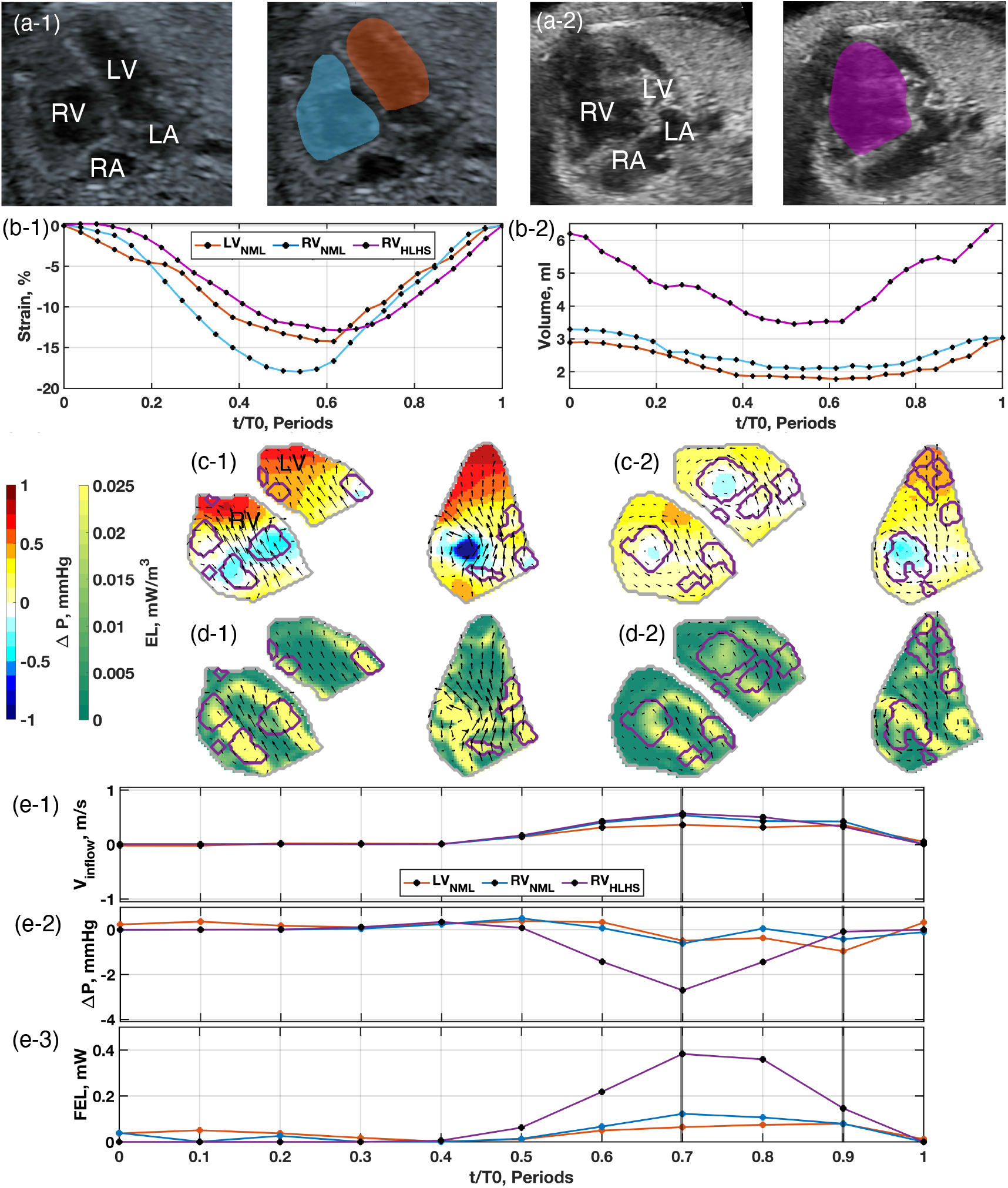
Comparison of echocardiographic measurements from a control heart and an HLHS heart at 33-weeks gestation. Control (NML; a-1) and HLHS (a-2) subject segmentations demonstrate identified boundary quality. Strain analysis (b-1) indicates comparable ventricular deformation in utero. Volume analysis (b-2) shows the HLHS RV is in overload. Peak diastolic pressure fields (c-1) show a large vortex develops along the free-wall of the HLHS RV, inducing greater energy loss (d-1). Late diastole pressure fields (c-2) and energy loss (d-2) behave similarly to early diastole. Timeseries curves are marked at early diastole and late diastole with gray lines. Inflow velocity (e-1) is comparable in utero. Intraventricular pressure difference (ΔP) (e-2) shows elevated pressure recovery for the HLHS heart. FEL measurements (e-3) show the HLHS heart has a two-fold increase in loss across the field due to the free-wall vortex.

### Systolic parameters in Postnatal HLHS RV

Measured parameters for the neonate HLHS hearts and controls are provided in Table 2. Major differences were observed for morphology as well as for systolic parameters. Observed volume changes occurred for both the HLHS RV and controls, but separation of SV and CO from the control LV and RV was not observed. HLHS RV s’ was significantly elevated compared to the control LV but not the RV. Peak GLS for the HLHS RV was significantly depressed compared to the control LV and RV.

### Diastolic Parameters in Postnatal HLHS RV

Major differences were observed for several of the same diastolic parameters for neonates’ hearts as fetal hearts. Comparable peak e’ (cm/s) was observed between the HLHS RV and the controls. HLHS RV E remained significantly elevated compared to the control RV and LV. HLHS RV E/e’ remained elevated compared to the control RV and LV. HLHS RV E/ increased compared to the control RV and LV. Peak FEL, peak VS, and, recovery Δ*P* each increased for the HLHS RV and control LV but remained unchanged for the control RV and were significantly different between conditions. AV-to-P_min_ remained unchanged for the HLHS RV control RV and LV but were significantly different between conditions.

### Qualitative assessment between postnatal HLHS RV

Representative changes in ventricular volumes, strains, and intracardiac flows are illustrated in Figure 3 for a neonate HLHS RV and a control. The major differences observed for the HLHS RV are more prevalent after birth. The increased volume and altered AV valve position for the HLHS RV produced an asymmetric vortex pair during diastole (Figure 3c, d) with the free wall vortex occupying a larger area than the septal wall vortex. An augmented pressure field was again observed during both diastole phases (Figure 2c-1, c-2), where the free wall vortex had stronger low pressure and the apex had stronger high pressure compared to the healthy heart. Stronger EL was observed both diastole phases (Figure 2d-1, d-2) compared to the healthy heart due to more disordered flow which produces greater shear. The HLHS RV timeseries (Figure 3c, d) did not showed fused diastole phases, stronger reversal IVPD and peak FEL compared to the healthy heart due to altered filling patterns.

**Figure 3:**
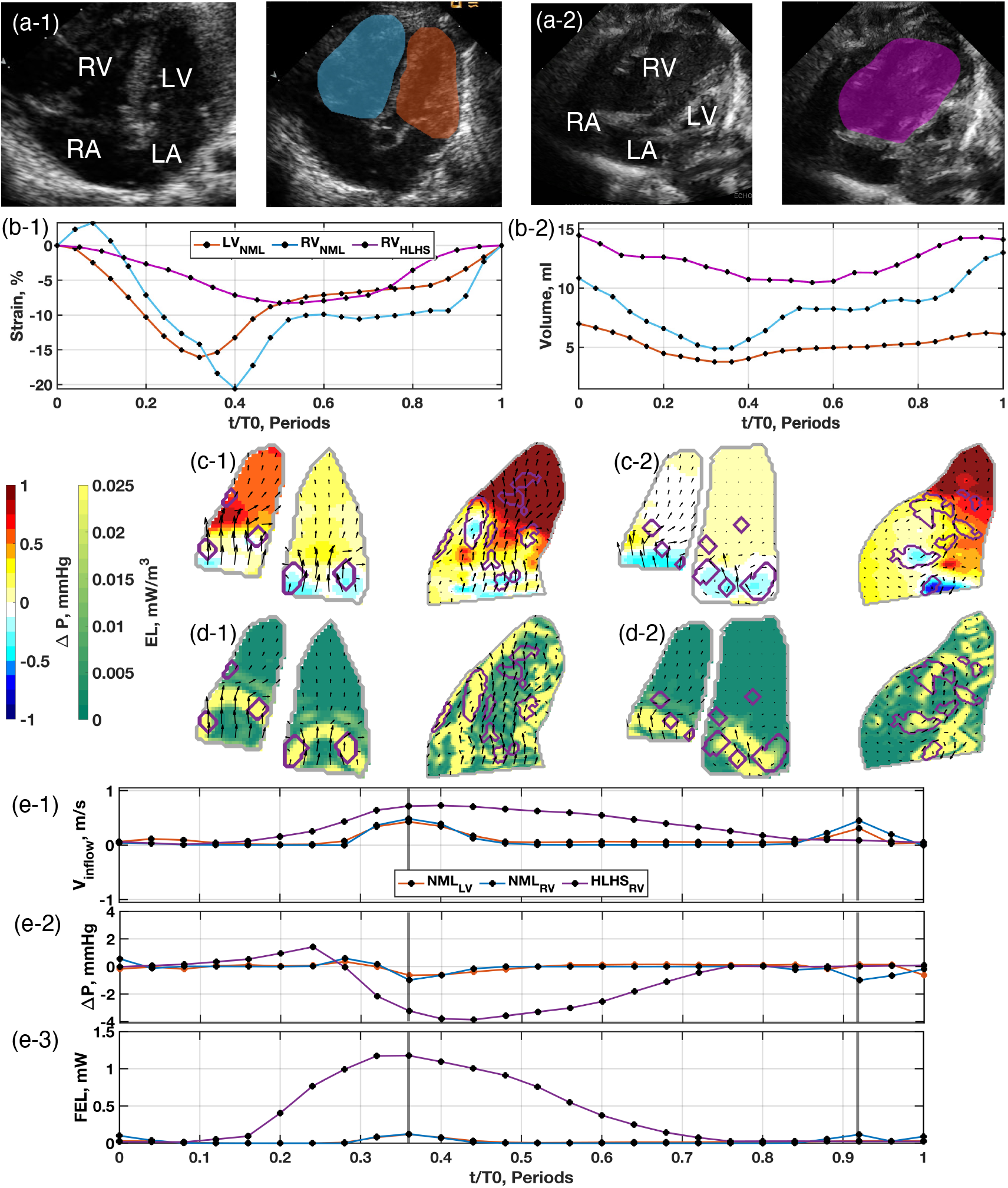
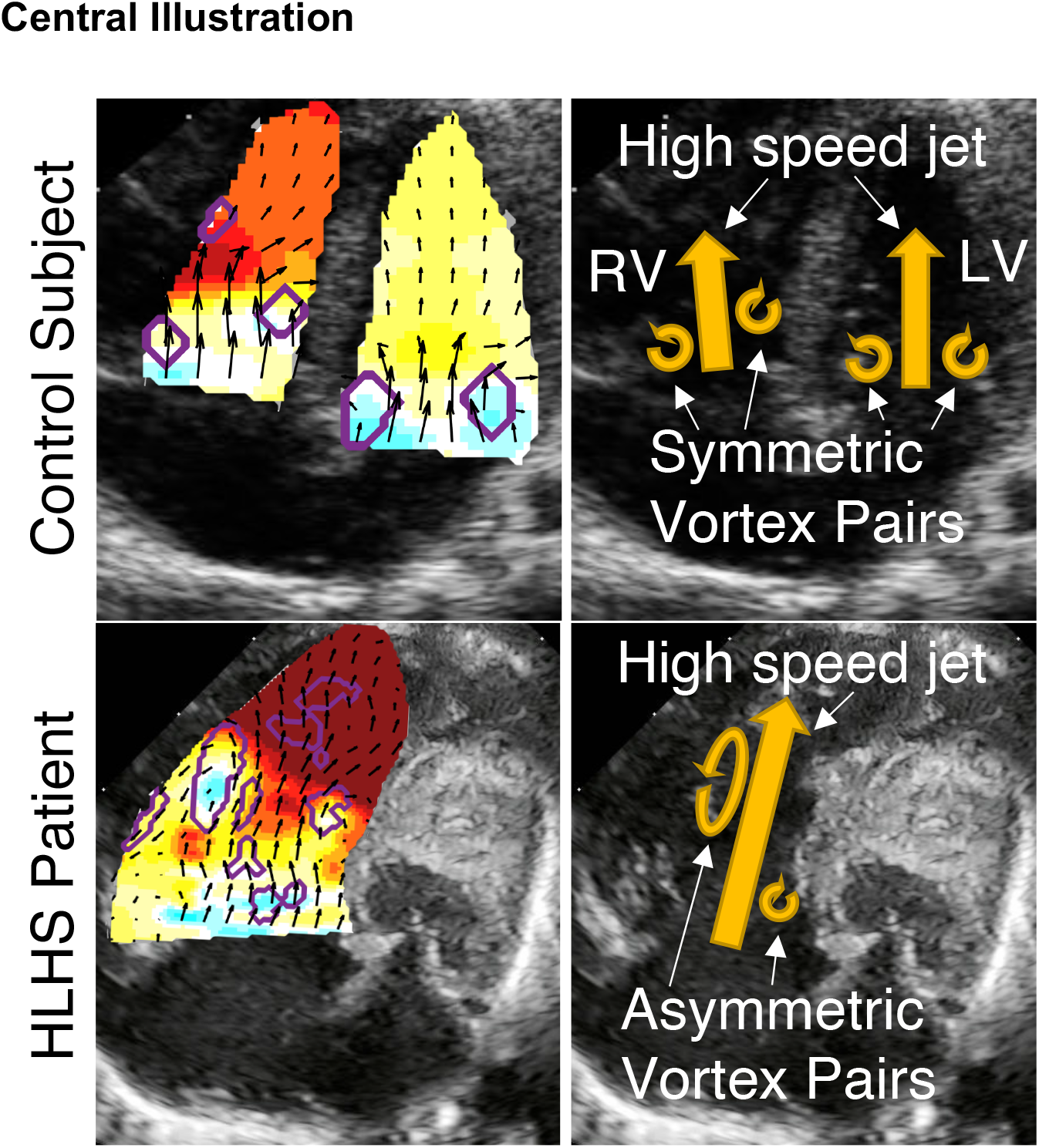
Comparison of echocardiographic measurements from a control heart and an HLHS heart at first week of birth. Control (NML; a-1) and HLHS (a-2) subject segmentations demonstrate identified boundary quality. Strain (b-1) is significantly reduced in the HLHS RV. Volume analysis (b-2) shows the HLHS RV is in overload. Peak diastolic pressure fields (c-1) show flow in the HLHS RV is disordered, inducing greater energy loss (d-1). Late diastole pressure fields (c-2) and energy loss (d-2) behave similarly to early diastole. Timeseries curves are marked at early diastole and late diastole with gray lines. Inflow velocity (e-1) is comparable in magnitude, but the HLHS RV inflow shows fused early and late diastole. Intraventricular pressure difference (ΔP) (e-2) shows elevated pressure recovery for the HLHS heart. FEL measurements (e-3) show the HLHS heart has a nearly ten-fold increase in loss across the field. Central Figure: Demonstration of the vortex pairs that form along the annular valve leaflet tips during diastolic filling. In the healthy heart (top row), the pair that forms is nearly symmetric and acts as liquid rollers, helping push blood toward the apex to wash out each chamber. In the HLHS patient (bottom), the pair is asymmetric, with a larger free wall vortex, which causes greater energy loss for proper filling and wash out of the chamber.

## Discussion

Assessing diastolic function non-invasively remains a critical challenge for HLHS patients. Even after successful surgical palliation HLHS patients can suffer from reduced exercise performance, decreased functional status, or require a heart transplant if diastolic dysfunction is present (10,11). However, unlike systolic function, conventional imaging measurements do not correlate well with diastolic function, particularly in the perinatal/infant period (22). Thus, the deterioration of diastolic function in HLHS patients is not understood, precluding the development of well-timed primary prevention strategies. This study quantified cardiac function biomechanics of normal and HLHS hearts from fetal and neonatal echocardiograms. We found that under HLHS RV remodeling, altered diastolic blood flow and reduced compliance associated with diastolic dysfunction (DD) can be observed *in* and *ex utero*.

HLHS defects are readily identified *in utero* with fetal ultrasound as the RV remodels to support the pulmonary and systemic circulations, reflected in the SV and CO parameters reported in Table 2. The combined SV between HLHS and control hearts closely matched prenatally (4.20 mL vs 4.37 mL), indicating the enlarged RV develops to support extra blood volume. While these parameters corroborate remodeling, they provide no information on diastolic function.

The additional quantitative results and qualitative analysis presented in this work enable us to make observations on diastolic function and the presence of DD in HLHS subjects. The E/e’ quantity for the HLHS RV of fetal (16.2) and neonates (16.2) subjects is above the healthy range (>12), associated with elevated filling pressures (16). This is coupled with elevated IVPD required to empty and fill the HLHS RV (Figure 2e-2, Figure 3e-2). Altered pressures are a result of increased stiffness and reduced contractility.

The early and late diastole phases were consistently fused in HLHS subjects (demonstrated in Figure 2e-1, Figure 3e-1), which is associated with DD. This fused pattern manifests because the HLHS RV is exposed to both systemic and pulmonary pressures. While less pronounced in utero, this altered filling pattern becomes much more apparent after birth with the onset of spontaneous respirations and predicted fall in pulmonary vascular resistance.

The increased HLHS RV volume allows for altered diastolic flow, exhibited in larger vortex formations (Figure 2c, Figure 3c), more disorganized flow (Figure 2d, Figure 3d), and greater flow energy loss (Figure 2d and Figure 3d). In healthy hearts, a donut-like vortex ring forms at the annular valve leaflet tips (23), which appears as two counter-rotating vortexes, as seen in the Central Figure. Normally, these vortices help aid in efficient ventricular filling (24). In the HLHS RV, the free wall vortex occupies a larger area because of the increased volume, resulting in greater flow energy loss which reduces efficient flow redirection prior to systolic ejection.

Prior studies observed that the fetal HLHS RV could suffer from diastolic dysfunction. In a 2008 study, Szwast et al reported that fetal HLHS hearts had diastolic dysfunction based on altered myocardial performance index (MPI), a ratio of the isovolumic times and ejection duration (25). Two separate studies have reported similar MPI findings as well (26,27). Brooks et al also reported altered late diastolic filling velocity and more reliance on atrial contraction. While these studies observed fetal HLHS heart diastolic dysfunction, they relied on imaging normally collected in fetal echo studies. This work further demonstrates that novel hydrodynamic parameters, which can be quantified from scans collected during anatomy ultrasounds, can also detect functional changes in the HLHS RV including the presence of diastolic dysfunction.

This study, for the first time, quantifies hydrodynamics parameters *in utero* and at birth, elucidating the differences in cardiac biomechanics between HLHS and normal hearts. Advancements in 4D MRI are improving the capabilities of the fetal heart assessment (28), but it has only recently been demonstrated in humans (29). Significant motion, lack of ECG gating, and low image resolution similarly affect 4D MRI. Importantly, while the above methods can measure novel parameters that better inform on cardiac function, they have been developed for adult populations, hence, they are not directly applicable to fetal and neonatal echocardiograms.

### Study Limitations

Our study cohort size (10 HLHS subjects) may not yet capture the statistics of the broader population. Imaging limitations in fetal echocardiography require novel measurement algorithms to be developed to ensure the most robust evaluation possible. Although the methods employed in this work have been demonstrated in prior studies, this is the first time the tools have been used to build a collective picture of fetal and neonatal biomechanics from echocardiography. Future work must be conducted to further verify the methods against the current gold standards in cardiac imaging. We will pursue additional fetal and pediatric measurements in future studies to enable further quantification of functional differences. Finally, the implemented analysis method described here is automated but does require three user-selected initialization points. A fully automated method will be explored, where the three user-selected points will be replaced with three points found by AI-based feature detection tools.

## Conclusions

This work evaluated cardiac function biomarkers for HLHS patients and age-matched controls from fetal and neonate echocardiograms. The methods used in this work collected conventional biomarkers routinely gathered during examination along with novel hemodynamic and hydrodynamic biomarkers derived from a new color Doppler reconstruction algorithm. Conventional biomarkers indicate that the HLHS RV contracts and deforms like functionally normal bi-ventricle hearts, even as SV and CO reflect the added volume taken on. It is only through observing the novel biomarkers that functional changes and detection of diastolic dysfunction in the HLHS RV can be observed. Importantly, these new biomarkers allow better quantification of myocardial performance, potentially improving the diagnosis and management of fetal heart failure. Altered hemodynamics and reduced ventricular relaxation were observed in the presence of a severe CHD, indicating the methods may provide earlier detection of anomalies *in utero* and lead to improving treatment practices *ex utero*.

## Perspectives

### Competency in Medical Knowledge

A challenge in the treatment and management of HLHS is that current echocardiographic metrics do not provide an adequate understanding of the RV to predict outcomes across the three stages of palliation. In this work, we have developed and tested novel biomechanics tools to provide easily obtained information on ventricular flow, energy loss, and tissue motion in fetal and neonatal hearts. These tools show significant differences between controls and HLHS. RV diastolic dysfunction begins in utero, and FEL is significant for HLHS RV performance. These tools can be used to model and test changes in pre-operative or operative management to improve outcomes in HLHS. Finally, longitudinal application of these analysis tools to children with complex congenital heart disease over time, such as HLHS, may allow the determination of ventricular characteristics that predict outcome, and may improve medical management.

### Translational Outlook

This study demonstrates the development and application of a novel set of integrated automated echocardiographic analysis tools to allow measurements of fetal and neonatal ventricular hemodynamics and function. These tools must next be prospectively validated on a larger cohort of subjects. When applied to patient care, this tool will allow earlier understanding and quantification of complex congenital heart diseases, such as HLHS, which may improve early management in this challenging group of high-risk infants and children. This will also advance our understanding and medical knowledge of fetal and neonatal ventricular function in complex heart disease.

## Data Availability

Data will be made available upon request.

## Abbreviations

AV: Atrioventricular
ALAX: Apical long axis
CFI: color flow imaging
CO: Cardiac output
EL: Energy loss
HLHS: Hypoplastic Left Heart Syndrome
VS: Vortex strength
SV: Stroke volume
Δ*P*: Pressure difference
IVPD: Intraventricular pressure difference

## Acknowledgements

*None*.

