## Appendix 1 for "Fetal and Neonatal Echocardiographic Analysis of Biomechanical Alterations for the Hypoplastic Left Heart"

### Appendix A

#### 1. Speckle Tracking Algorithm

Input atrioventricular valve (AV) annulus ends at the septal and lateral walls positions and the ventricle apex position are tracked temporally using Fourier domain cross-correlation (1, 2). Between sequential frames a subwindow of pixels centered around each position is taken. Each spatial subwindow pair, $w_{1}\left( x,y \right)$ and $w_{2}\left( x,y \right)$, is converted to the Fourier domain via the Fast Fourier Transform (FFT),

$$G_{i}\left( u,v \right)\mathcal{=F}\left( w_{i}\left( x,y \right) \right).$$

Equation 1

The Fourier Transforms for each pair are subsequently cross-correlated,

$$R\left( x,y \right)=\mathcal{F}^{-1}\left( G_{1}\left( u,v \right)\bar{G}_{2}\left( u,v \right) \right),$$

Equation 2

where $\mathcal{F}^{-1}$ is the inverse Fourier Transform (FT), $\left( u,v \right)$ are wavenumbers proportional to spatial coordinates $\left( x,y \right),$ and the overbar symbolizes the complex conjugate. The displacement between the pair is computed by the peak location of the cross-correlation $R$

$$\left( \Delta x,\Delta y \right)={argmax}_{\left( x,y \right)}\left( R \right),$$

Equation 3

Each position is updated by adding the displacements to the current positions, which are then used as new centers for the next subwindow pair in the temporal image sequence.

#### 2. Global Longitudinal Strain Algorithm

For each pair of co-registered frames containing the ventricle, a specialized correlation kernel is used to estimate GLSr between frames. The following steps are performed to calculate GLSr using the specialized kernel. First, the FFT for each frame in the pair is computed based on Equation 1. Next, the magnitudes of each FT are calculated, removing the phase content. Subsequently an interpolation is performed to render the FT magnitudes on a logarithm scale. Each re-scaled FT magnitude is then quartered into new subimages and their FFT is computed. The phase-filtered cross-correlation is then calculated for each subimage pair, like Equation 2, and their results are ensemble averaged. Finally, the displacements are found from cross-correlation peak based on Equation 3, which are converted from the logarithm scale to the image scale, returning a measurement for longitudinal strain. Each step is shown in Figure 1 of (4).

#### 3. Unsupervised Segmentation Algorithm

Identifying the ventricle boundary with the unsupervised segmentation algorithm occurs in four steps: (1) image pre-processing, (2) path-finding with Dijkstra's algorithm using an echocardiogram specific cost-matrix, (3) post-processing, and (4) volume calculation.

Image pre-processing works to normalize the contrast-to-noise ratio (CNR), smooth added image noise, and transform the image onto a polar grid. These steps improve the reliability of the Dijkstra's algorithm in segmenting the boundary. Image CNR normalization uses adaptive contrast enhancement, which is computed for each image pixel,

$${Im}_{ACE}\left( x,y \right)=\frac{Im\left( x,y \right)-{Im}_{avg}}{{Im}_{max}-{Im}_{avg}}.$$

Equation 4

In Equation 4, ${Im}_{ACE}$ is the ACE pixel intensity, $Im$ is the raw pixel intensity, ${Im}_{avg}$ is the average pixel intensity of a line from the AV valve ends to the ventricle apex, and ${Im}_{max}$ is the maximum intensity in the entire image. Median filtering with an 11 x 11 pixel window is used to smooth the noise. The polar transformation, last in the image pre-processing, warps the image from a Cartesian grid $\left( x,y \right)$ to a polar grid $\left( \rho,\theta\right)$ relative to the center of the ventricle $\left( x_{c},y_{c} \right)$, using the equations,

$$\rho=\sqrt{\left( x-x_{c} \right)^{2}+\left( y-y_{c} \right)^{2}},\theta=atan\left( \frac{y}{x} \right).$$

Equation 5

By transforming the ventricle image, the curve or path-finding of the ventricle simplifies the search for a curve that connects the two AV valve nodes through the ventricle apex. A cost-matrix for the performing the Dijkstra's algorithm is constructed by initially treating each pixel as a *graph-node*. A peak prominence- -based cost was chosen; in this approach along each line of $\theta$ the local peak intensities $\left( \hat{I} \right)$ and their height compared to the local minimum $\hat{\left( P \right)}$ are computed, which are used in the cost equation $\left( C \right)$

$$C=15\left( \hat{P}+\hat{I} \right)^{-1}.$$

Equation 6

In order to reduce computation time, nodes larger than 1.25 the mean of $C$ are discarded. The Dijkstra's algorithm (3) was used iteratively, allowing the method to detect the lowest cost for a limited number of lines of $\theta$. This step size was varied from 2^o^ to 14^o^ in steps of 2^o^. The seven curves generated from the iterative processing were fit with a smoothing-spline radial basis function to produce a single representative curve, or the ventricle segmentation. This segmentation is converted back onto a Cartesian grid.

Each curve is post-processed with a boundary smoothing tool to ensure the segmentation appears physiologically consistent. This post-processing finds all possible paths within a neighborhood of nodes in the curve, evaluates the gradients for all paths and the current path, selects the path with the smallest gradient. Two iterations are performed to ensure no over-smoothing occurs.

#### 4. Doppler Vector Reconstruction Algorithm

The color Doppler vector reconstruction algorithm used in this work is based on the relationship between the flow rate and fluid rotation. The algorithm processes a single-color Doppler frame by (1) converting the color signal to velocities, (2) segmenting the ventricle from the gray-scale signal and imposing boundary conditions, and (3) using a numerical solver to reconstruct the velocity field.

For each color Doppler recording, the user inputs the AV annulus end positions and ventricle apex position from the first recorded frame like the algorithms above. The user will also crop the velocity color scale from the first recorded frame and provide its upper and lower velocity limits, in centimeters per second. A velocity range the same length as the color velocity scale is constructed.

A single frame is first separated into a color-signal image and a gray-scale image. This step avoids adding noise in further calculations and ensures only the blood flow signal is used in the reconstruction. Each pixel in the color-signal is converted to a velocity by converting its color value on the image velocity color scale to a velocity on the constructed velocity range. The output is a map of velocity measurements that correspond to the color-signal of the current color Doppler frame.

The gray-scale image is evaluated using the unsupervised segmentation algorithm and the current AV annulus end positions and apex position to obtain a segmentation curve. This curve is converted to a binary mask with the same dimensions as the map of velocity measurements. Velocity measurements along a line between the AV annulus end positions are used to impose inflow/outflow conditions for the solver,

$$\psi_{flow}\left( s \right)=\int_{s_{0}}^{s_{a}} u\left( s \right)d\xi+\psi_{0}.$$

Equation 7

In equation 7, $\psi_{0}$ is the initial flow rate which we set to zero, $\psi_{flow}$ is the flow rate a location along the line $s$, and $s_{0}$ and $s_{a}$ are the annulus end positions of the line. A free-penetration condition is assumed along the ventricle walls to ensure flow rate is balanced, which is assumed to be linearly distributed,

$$\psi_{wall}\left( s \right)=f\left( s,u \right).$$

Equation 8

The Doppler Vector Reconstruction (DoVeR) solver is used to reconstruct the 2D velocity vector field (5) using the boundary conditions and map of velocities. An initial fluid rotation source term, $\omega^{0}$, is quantified. An initial error is provided, set to be infinite. DoVeR is an iterative reconstruction tool, so the error is checked against a threshold for each iteration; this threshold was set to 10^-8^. The error is computed from the L2-norm of the difference between the current and previous iteration reconstructed vector fields,

$$\varepsilon=\frac{\left\| \mathbf{u}^{n}\boldsymbol{-}\mathbf{u}^{n-1} \right\|}{\left\| \mathbf{u}^{n} \right\|}.$$

Equation 9

So long as error is not minimized the iteration number, $n$, is increased and new flow rates through the ventricle $\psi^{n}$ are computed from the fluid rotation $\omega^{n-1}$ by LU-decomposition of,

$$\ddot{\boldsymbol{D}}\psi^{n}=-\omega^{n-1}\boldsymbol{,}$$

Equation 10

where $\ddot{\boldsymbol{D}}$ is the second-order derivative operator of size $N\times N$ with a 3-point stencil size, and $\omega^{n-1}$ and $\psi^{n}$ are $N\times1$ vectors. The quantity *N* is the total number of points within the ventricle boundary. The reconstructed 2D velocity vector field, $\mathbf{u}^{n}$, is computed with,

$$\mathbf{u}^{\boldsymbol{n}}\boldsymbol{=}\left[ \begin{matrix} u_{x}^{n} & u_{y}^{n} \end{matrix} \right]\boldsymbol{=}\left[ \begin{matrix} {\dot{\boldsymbol{D}}}_{\boldsymbol{y}}\psi^{n} & {\dot{\boldsymbol{D}}}_{\boldsymbol{x}}\psi^{n} \end{matrix} \right]\boldsymbol{,}$$

Equation 11

where ${\dot{\boldsymbol{D}}}_{\boldsymbol{x}}$ and ${\dot{\boldsymbol{D}}}_{\boldsymbol{y}}$ are first-order derivative operators of size $N\times N$ with 3-point stencil size for derivatives in *x* and *y.* All non-zero velocities from the input velocity map must remain constant in $u_{y}^{n}$to further constrain the solver. Zero velocities from the input velocity map can be replaced but are threshold to ±10% of the maximum of the input velocity map. Finally, the fluid rotation $\omega^{n}$ is updated using the discrete formulation,

$\omega^{n}\boldsymbol{=}{\dot{\boldsymbol{D}}}_{\boldsymbol{x}}u_{y}^{n}\boldsymbol{-}{\dot{\boldsymbol{D}}}_{\boldsymbol{y}}u_{x}^{n}$.

Equation 12

##### 5. Reconstruction post-processing

Post-processing the reconstructed 2D vector fields helps quantify ventricle flow efficiency. The vector fields are quantified using finite-differencing. Flow energy loss (EL) is calculated by,

$$EL=\int\left[ {2\left( \frac{\partial u_{x}}{\partial x} \right)}^{2}+{2\left( \frac{\partial u_{y}}{\partial y} \right)}^{2}+\left( \frac{\partial u_{x}}{\partial y}+\frac{\partial u_{y}}{\partial x} \right)^{2} \right]dV,$$

Equation 13

where $V$ is the volume of the region of flow, found by multiplying the 2D area of the ventricle by the AV valve diameter. Vortices are identified based on flow rotation, using the λ_CI_ criterion (6). The total vortex strength (VS) from this algorithm is,

$$VS=\sum\left| \int\omega dA \right|_{vortex},$$

Equation 14

where $A$ is the area of each identified vortex. The subscript indicates summation across all identified vortex structures. The 2D pressure field, $P$, inside the ventricles is computed using the pressure Poisson equation,

$$\nabla^{2}P=\left( \frac{\partial u_{x}}{\partial x} \right)^{2}+\left( \frac{\partial u_{y}}{\partial y} \right)^{2}+2\left( \frac{\partial u_{x}}{\partial y}\frac{\partial u_{y}}{\partial x} \right).$$

Equation 15

Neumann boundary conditions account for temporal variation of pressure. The solver reconstructs $P$ using a least squares formulation, through an in-house algorithm described previously in (7). This analysis returns quantities for peak early and late AV valve velocities (E,A), EL, VS, and peak suction and reversal AV valve-to-apex pressure differences $\left( \Delta P \right)$.

### Appendix B

| Quantity | Abbreviation | Definition |
| --- | --- | --- |
| Cardiac Output | CO | Volume flow rate the heart circulates through the body in one minute |
| Stroke Volume | SV | Blood volume pumped out of the ventricles during systole |
| Peak early diastolic  annular velocity | e’ | Maximum velocity the annular plane that holds the AV valve moves during suction filling phase as the ventricle relaxes |
| Peak late diastolic  annular velocity | a’ | Maximum velocity the annular plane that holds the AV valve moves during atrial contraction filling phase |
| Peak systolic  annular velocity | s’ | Maximum velocity the annular plane that holds the AV valve moves as the ventricle contracts |
| Peak early AV  filling velocity | E | Maximum velocity of blood flowing through the AV valve during suction filling phase as the heart relaxes |
| Peak late AV  filling velocity | E | Maximum velocity of blood flowing through the AV valve during atrial contraction phase |
|  | E/e’ | Ratio for the blood flow velocity to the annular plane velocity during the suction filling phase; correlates to filling pressure |
|  | E/e’ | Ratio for the suction filling blood flow velocity to the atrial contraction blood flow velocity |
| Peak global  longitudinal strain | \|GLS\|_max_ | A percentage change in length of the ventricle from the start of systole to the start of diastole |
| Peak systolic GLS  rate | GLSrs | Maximum rate of change of the ventricle length during systole |
| Peak diastolic GLS  rate | GLSre | Maximum rate of change of the ventricle length during diastole |
| Flow energy loss | FEL | Maximum of total energy dissipated by viscous losses over the ventricular volume during diastole |
| Viscous Strength | VS | Maximum of total rotation over the ventricle volume during diastole |
| Intraventricular pressure difference | ΔP | Difference in pressure between the atrioventricular valve and apex |
|  | Suction ΔP | IVPD at the beginning of diastole that helps initiate filling |
|  | Recovery ΔP | IVPD at peak diastole when the flow stops accelerating |
|  | Ejection ΔP | IVPD at peak systole when blood flow rate is highest along the outflow tracts |
| Minimum pressure location | Min. ΔP Location | Spatial location of minimum pressure inside the ventricle during diastole |

### Appendix C

|  |  | HLHS RV (n = 10) | CTRL LV (n = 12) | | CTRL RV (n = 12) | |
| --- | --- | --- | --- | --- | --- | --- |
|  |  | $\mu\pm1\sigma$ | $\mu\pm1\sigma$ | p-value | $\mu\pm1\sigma$ | p-value |
| Stroke Volume | Prenatal | $4.20\pm2.08$ | $1.85\pm0.79$ | **0.002** | $2.52\pm1.18$ | **0.030** |
| (ml) | Postnatal | $4.38\pm2.54$ | $3.23\pm1.22$ | 0.187 | $3.42\pm3.05$ | 0.455 |
|  | p-value | 0.868 | **0.004** |  | 0.366 |  |
| Cardiac Output | Prenatal | $618\pm324$ | $282\pm111$ | **0.004** | $386\pm183$ | *0.052* |
| (ml/min) | Postnatal | $675\pm530$ | $426\pm181$ | 0.144 | $444\pm429$ | 0.283 |
|  | p-value | 0.771 | **0.034** |  | 0.683 |  |
| s’, | Prenatal | $2.24\pm1.57$ | $2.77\pm1.02$ | 0.365 | $3.37\pm1.65$ | 0.117 |
| (cm/s) | Postnatal | $3.17\pm1.01$ | $2.20\pm0.81$ | **0.021** | $3.00\pm0.87$ | 0.677 |
|  | p-value | 0.131 | 0.149 |  | 0.498 |  |
| e’ | Prenatal | $3.63\pm1.96$ | $3.45\pm1.07$ | 0.798 | $4.68\pm2.16$ | 0.246 |
| (cm/s) | Postnatal | $4.22\pm1.32$ | $3.59\pm1.85$ | 0.374 | $3.93\pm1.53$ | 0.639 |
|  | p-value | 0.429 | 0.836 |  | 0.345 |  |
| a’ | Prenatal | $2.16\pm1.48$ | $2.50\pm1.23$ | 0.568 | $3.47\pm2.42$ | 0.157 |
| (cm/s) | Postnatal | $3.15\pm1.68$ | $1.35\pm0.76$ | **0.013** | $1.98\pm0.62$ | **0.045** |
|  | p-value | 0.180 | **0.032** |  | *0.063* |  |
| E | Prenatal | $42.1\pm10.0$ | $30.4\pm6.5$ | **0.004** | $35.0\pm9.6$ | 0.105 |
| (cm/s) | Postnatal | $62.0\pm9.4$ | $42.5\pm17.5$ | **0.006** | $34.6\pm16.8$ | **< 0.001** |
|  | p-value | **< 0.001** | **0.045** |  | 0.950 |  |
| A | Prenatal | $32.7\pm11.5$ | $22.8\pm6.9$ | **0.024** | $22.4\pm12.8$ | **0.032** |
| (cm/s) | Postnatal | $40.8\pm12.2$ | $35.1\pm11.4$ | 0.294 | $31.3\pm14.2$ | 0.128 |
|  | p-value | 0.132 | **0.007** |  | *0.086* |  |
| E/e’ | Prenatal | $16.2\pm11.3$ | $10.0\pm4.6$ | 0.108 | $9.5\pm6.2$ | *0.097* |
|  | Postnatal | $16.2\pm5.8$ | $12.2\pm5.4$ | 0.122 | $10.1\pm3.4$ | **0.011** |
|  | p-value | 0.998 | 0.338 |  | 0.765 |  |
| E/A | Prenatal | $1.27\pm0.39$ | $1.36\pm0.44$ | 0.657 | $1.27\pm0.55$ | 0.962 |
|  | Postnatal | $1.61\pm0.44$ | $1.40\pm0.65$ | 0.397 | $0.84\pm0.27$ | **< 0.001** |
|  | p-value | *0.082* | 0.859 |  | *0.066* |  |
| \|GLS\|_max_ | Prenatal | $19.4\pm6.3$ | $15.5\pm3.0$ | *0.083* | $19.2\pm10.0$ | 0.966 |
| (%) | Postnatal | $16.6\pm3.6$ | $19.3\pm5.2$ | 0.183 | $23.6\pm8.2$ | **0.023** |
|  | p-value | 0.249 | **0.045** |  | 0.265 |  |
| GLSrs | Prenatal | $1.92\pm0.33$ | $2.05\pm0.68$ | **0.011** | $1.97\pm0.58$ | **0.013** |
| (s^-1^) | Postnatal | $1.30\pm0.30$ | $1.51\pm0.34$ | 0.133 | $1.89\pm0.48$ | **0.003** |
|  | p-value | 0.382 | **0.023** |  | 0.722 |  |
| GLSre | Prenatal | $1.82\pm1.33$ | $1.27\pm0.89$ | 0.296 | $1.47\pm0.99$ | 0.504 |
| (s^-1^) | Postnatal | $0.91\pm0.58$ | $1.53\pm0.52$ | **0.016** | $1.83\pm1.15$ | **0.034** |
|  | p-value | *0.064* | 0.412 |  | 0.430 |  |
| Flow energy loss | Prenatal | $0.25\pm0.22$ | $0.09\pm0.07$ | **0.026** | $0.14\pm0.11$ | 0.149 |
| (mW) | Postnatal | $0.56\pm0.49$ | $0.19\pm0.20$ | **0.044** | $0.16\pm0.19$ | **0.030** |
|  | p-value | *0.077* | 0.103 |  | 0.756 |  |
| Vortex Strength | Prenatal | $241\pm108$ | $104\pm42$ | **< 0.001** | $146\pm55$ | **0.017** |
| (cm^2^/s) | Postnatal | $322\pm135$ | $197\pm110$ | **0.035** | $155\pm110$ | **0.007** |
|  | p-value | 0.143 | **0.016** |  | 0.826 |  |
| Suction ΔP | Prenatal | $0.23\pm0.24$ | $0.23\pm0.11$ | 0.96 | $0.19\pm0.15$ | 0.60 |
| (mmHg) | Postnatal | $0.79\pm0.71$ | $0.39\pm0.30$ | 0.13 | $0.46\pm0.40$ | 0.25 |
|  | p-value | **0.03** | 0.13 |  | **0.05** |  |
| Recovery ΔP | Prenatal | $-1.17\pm0.81$ | $-0.50\pm0.27$ | **0.018** | $-0.61\pm0.22$ | **0.039** |
| (mmHg) | Postnatal | $-2.34\pm1.13$ | $-1.10\pm1.02$ | **0.019** | $-0.68\pm0.63$ | **< 0.001** |
|  | p-value | **0.013** | *0.075* |  | 0.707 |  |
| Ejection ΔP | Prenatal | $0.14\pm0.21$ | $0.23\pm0.21$ | 0.292 | $0.16\pm0.17$ | 0.752 |
| (mmHg) | Postnatal | $0.38\pm0.30$ | $0.50\pm0.47$ | 0.483 | $0.34\pm0.47$ | 0.821 |
|  | p-value | **0.047** | *0.010* |  | 0.269 |  |
| Min. ΔP Location | Prenatal | $9.1\pm3.6$ | $4.6\pm1.9$ | **0.002** | $4.6\pm1.7$ | **0.001** |
| (mm) | Postnatal | $7.9\pm2.8$ | $3.6\pm2.0$ | **< 0.001** | $4.2\pm2.1$ | **0.003** |
|  | p-value | 0.419 | 0.254 |  | 0.599 |  |
